# DengueSeq: A pan-serotype whole genome amplicon sequencing protocol for dengue virus

**DOI:** 10.1101/2023.10.13.23296997

**Authors:** Chantal B.F. Vogels, Verity Hill, Mallery I. Breban, Chrispin Chaguza, Lauren M. Paul, Afeez Sodeinde, Emma Taylor-Salmon, Isabel M. Ott, Mary E. Petrone, Dennis Dijk, Marcel Jonges, Matthijs R.A. Welkers, Timothy Locksmith, Yibo Dong, Namratha Tarigopula, Omer Tekin, Sarah Schmedes, Sylvia Bunch, Natalia Cano, Rayah Jaber, Charles Panzera, Ian Stryker, Julieta Vergara, Rebecca Zimler, Edgar Kopp, Lea Heberlein, Andrea M. Morrison, Scott F. Michael, Nathan D. Grubaugh

**Affiliations:** Department of Epidemiology of Microbial Diseases, Yale School of Public Health, New Haven, Connecticut, United States of America; Yale Institute for Global Health, Yale University, New Haven, Connecticut, United States of America; Department of Biological Sciences, College of Arts and Sciences, Florida Gulf Coast University, Fort Myers, Florida, United States of America; Department of Pediatrics, Yale School of Medicine, New Haven, Connecticut, United States of America; Sydney Institute for Infectious Diseases, School of Medical Sciences, University of Sydney, NSW, Australia; Department of Medical Microbiology & Infection Prevention, Amsterdam UMC location AMC, Amsterdam, The Netherlands; Department of Infectious Diseases, Public Health Service of Amsterdam, Amsterdam, The Netherlands; Bureau of Public Health Laboratories, Division of Disease Control and Health Protection, Florida Department of Health, Tampa, FL, United States of America; Bureau of Public Health Laboratories, Division of Disease Control and Health Protection, Florida Department of Health, Jacksonville, FL, United States of America; Bureau of Epidemiology, Division of Disease Control and Health Protection, Florida Department of Health, Tallahassee, FL, United States of America; Department of Ecology and Evolutionary Biology, Yale University, New Haven, Connecticut, United States of America; Public Health Modeling Unit, Yale School of Public Health, New Haven, Connecticut, United States of America

**Keywords:** Genomic surveillance, next-generation sequencing, amplicon sequencing, whole-genome sequencing, dengue virus

## Abstract

**Background:** The increasing burden of dengue virus on public health due to more explosive and frequent outbreaks highlights the need for improved surveillance and control. Genomic surveillance of dengue virus not only provides important insights into the emergence and spread of genetically diverse serotypes and genotypes, but it is also critical to monitor the effectiveness of newly implemented control strategies. Here, we present DengueSeq, an amplicon sequencing protocol, which enables whole-genome sequencing of all four dengue virus serotypes.

**Results:** We developed primer schemes for the four dengue virus serotypes, which can be combined into a pan-serotype approach. We validated both approaches using genetically diverse virus stocks and clinical specimens that contained a range of virus copies. High genome coverage (>95%) was achieved for all genotypes, except DENV2 (genotype VI) and DENV 4 (genotype IV) sylvatics, with similar performance of the serotype-specific and pan-serotype approaches. The limit of detection to reach 70% coverage was 10^1^-10^2^ RNA copies/μL for all four serotypes, which is similar to other commonly used primer schemes. DengueSeq facilitates the sequencing of samples without known serotypes, allows the detection of multiple serotypes in the same sample, and can be used with a variety of library prep kits and sequencing instruments.

**Conclusions:** DengueSeq was systematically evaluated with virus stocks and clinical specimens spanning the genetic diversity within each of the four dengue virus serotypes. The primer schemes can be plugged into existing amplicon sequencing workflows to facilitate the global need for expanded dengue virus genomic surveillance.

## Background

The global burden of the mosquito-borne dengue virus continues to increase with an estimated 3.9 billion people at risk of infection [1]. During the last two decades, the number of reported dengue cases has increased by more than eightfold to over 4 million in 2019, particularly in tropical and sub-tropical regions in the Americas and Asia [2]. As of July 2023, close to 3 million dengue cases have been reported from the Americas this year, which has already surpassed the total number of 2.8 million cases reported in 2022 [3]. With global dengue cases surging, there is a critical need for genomic surveillance to track the emergence and spread of dengue virus lineages. These lineages consist of four antigenically distinct serotypes (DENV1-4) which can be further subdivided into genetically diverse genotypes [4,5]. Additionally, the rollout of novel vaccines and other biological interventions, such as the release of mosquitoes carrying the virus-inhibiting *Wolbachia* bacterium, warrant close monitoring of changes in dengue virus genetic diversity that may impact or reflect the effectiveness of these strategies [6,7].

Amplicon-based sequencing is a cost-effective, sensitive, and high-throughput method to conduct routine virus genomic surveillance. During the 2015-2016 Zika epidemic, challenges with sequencing the often low-titer virus using metagenomic protocols led to the development of PrimalSeq. PrimalSeq is a multiplexed tiled primer approach where overlapping amplicons are split into two PCR reactions [8,9]. Later, this approach was adapted as the primary sequencing method for SARS-CoV-2 (i.e., the “ARTIC” protocol) [10]. During the COVID-19 pandemic, investments in genomics infrastructure have significantly increased sequencing capacity globally, leading to a record number of over 15 million publicly available SARS-CoV-2 genomes. In contrast, less than 13,000 near-complete dengue virus genomes are publicly available on GenBank, and of those, only ∼20% were collected in the last five years [11,12]. Whole genome sequencing, rather than sequencing of targeted genes, is required to provide sufficient resolution for phylogenetic analyses, especially with more slowly evolving viruses such as dengue [13]. Moreover, a pan-serotype approach is needed to facilitate the sequencing of any serotype without the need for prior serotyping. Thus, designing a unified pan-serotype approach to sequence the global diversity of dengue virus, which fits within the established SARS-CoV-2 workflows, could rapidly accelerate the implementation of dengue virus genomic surveillance systems [14].

Here, we describe the development and validation of DengueSeq to sequence the known diversity of dengue virus from clinical samples. We designed primer schemes for all four dengue virus serotypes, and the serotype-specific primers can be used individually or combined as a universal pan-serotype amplicon-based sequencing approach. By adapting DengueSeq to currently existing amplicon-based sequencing workflows, our method can enhance the global capacity for whole-genome dengue virus sequencing.

## Results

### Primer scheme design

The PrimalSeq protocol uses a series of overlapping PCR amplicons generated with primers designed using PrimalScheme (https://primalscheme.com) to efficiently sequence virus genomes directly from clinical specimens [8,9]. Here, we used PrimalScheme to develop primer schemes for all four serotypes of dengue virus, which can be used with the amplicon-based sequencing protocols widely established during the COVID-19 pandemic [14]. We selected genetically diverse genomes within each of the dengue virus serotypes and then used PrimalScheme to generate four serotype-specific primer schemes (**Fig. 1A**). We chose genomes from all of the defined genotypes to ensure that the primer schemes captured the genetic diversity within each of the dengue virus serotypes (**Fig. 1B**). Specifically, we focused primarily on including viruses that have caused human outbreaks, and excluded genotypes with only incomplete genomes available and highly divergent genotypes (e.g. DENV1 genotype II, DENV2 genotype IV and VI (sylvatic), and DENV4 genotypes III and IV (sylvatic)). Each serotype primer scheme consists of 35-37 primer pairs with an average amplicon length of 400 bp (the default PrimalScheme amplicon length [8]). We also developed a bioinformatics pipeline that aligns sequencing reads against six default reference genomes (DENV1-4, and DENV2 and DENV4 sylvatics) through an iterative (non-competitive) loop to generate consensus genomes using iVar (**Fig. S1;** (https://github.com/grubaughlab/DENV_pipeline/tree/v1.0). Our assay, called DengueSeq, can be used individually with any of the four serotype-specific primer sets or combined into a pan-serotype approach, as validated below.

**Fig. 1:**
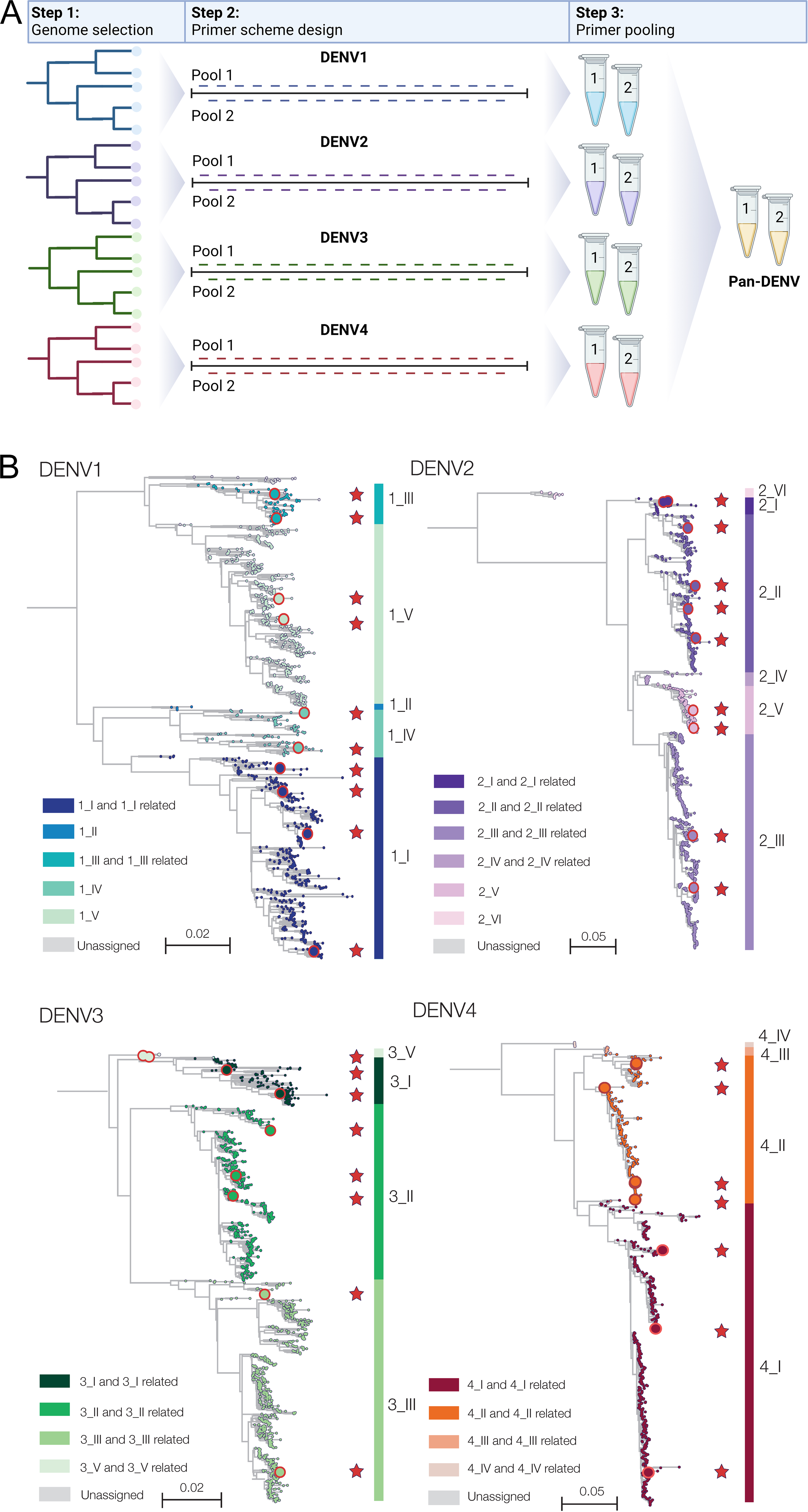
Overview of the DengueSeq primer design. A) For each of the four serotypes, a selection of genetically diverse genomes was made as input for PrimalScheme. Using PrimalScheme, four serotype-specific primer schemes were generated. Each scheme consists of 35-37 primer pairs which are split into two pools. Pan-serotype primer pools can be created by combining the serotype-specific pools 1 and 2, with which all four serotypes can be sequenced. Figure created with BioRender.com. B) Dengue virus genomes representing the genotypes within each of the four serotypes used to design the primer schemes. Red circles and stars indicate the genomes used to design the primer schemes for each serotype.

To account for high virus genetic diversity, we tested whether including degenerate nucleotides in primers at positions with mismatches to some viruses would help amplify a wider range of genetic diversity. For example, if some viruses had an adenine (A) at a primer position, and others had a guanine (G), then designing a primer with an “R” code would direct for some primers to be randomly synthesized with either an A or G nucleotide at that position. To make the alternate design, we aligned the primer sequences to all of the publicly available dengue virus genomes, and we included degenerate nucleotides at positions with alternate nucleotide frequencies above 10%. After comparing the “non-degenerate (original)” and “degenerate” DENV2 primer schemes, we found similar genome coverage when sequencing undiluted and diluted virus stocks (**Fig. S2**). As we found minimal differences between the two designs, we proceeded with further validation of the “non-degenerate” DENV1-4 primer schemes.

### Serotype-specific primer scheme validation

To perform our initial validation of the individual serotype-specific DengueSeq primer schemes, we sequenced 127 dengue virus stocks representing all of the defined genotypes that have caused human outbreaks (**Fig. 2**, **Fig. S3**). Based on results from the CDC dengue multiplex RT-qPCR assay [15], we sequenced undiluted RNA using the corresponding serotype-specific primer scheme and the Illumina COVIDSeq test. We used our bioinformatics pipeline to generate consensus genomes at a minimum depth of coverage of 20x and Genome Detective to assign genotypes [16]. We generated genomes with high genome coverage (>95%) in the coding sequence for all genotypes (**Fig. 2A, C, E, G**), except for the sylvatic genotypes (DENV2 genotype VI (60.0-91.2%) and DENV4 genotype IV (93.3-94.2%); **Fig. 2C, G**) that were not included in the primer design. These findings demonstrated that our serotype-specific primer schemes captures the genetic diversity present within each of the serotypes.

**Fig 2:**
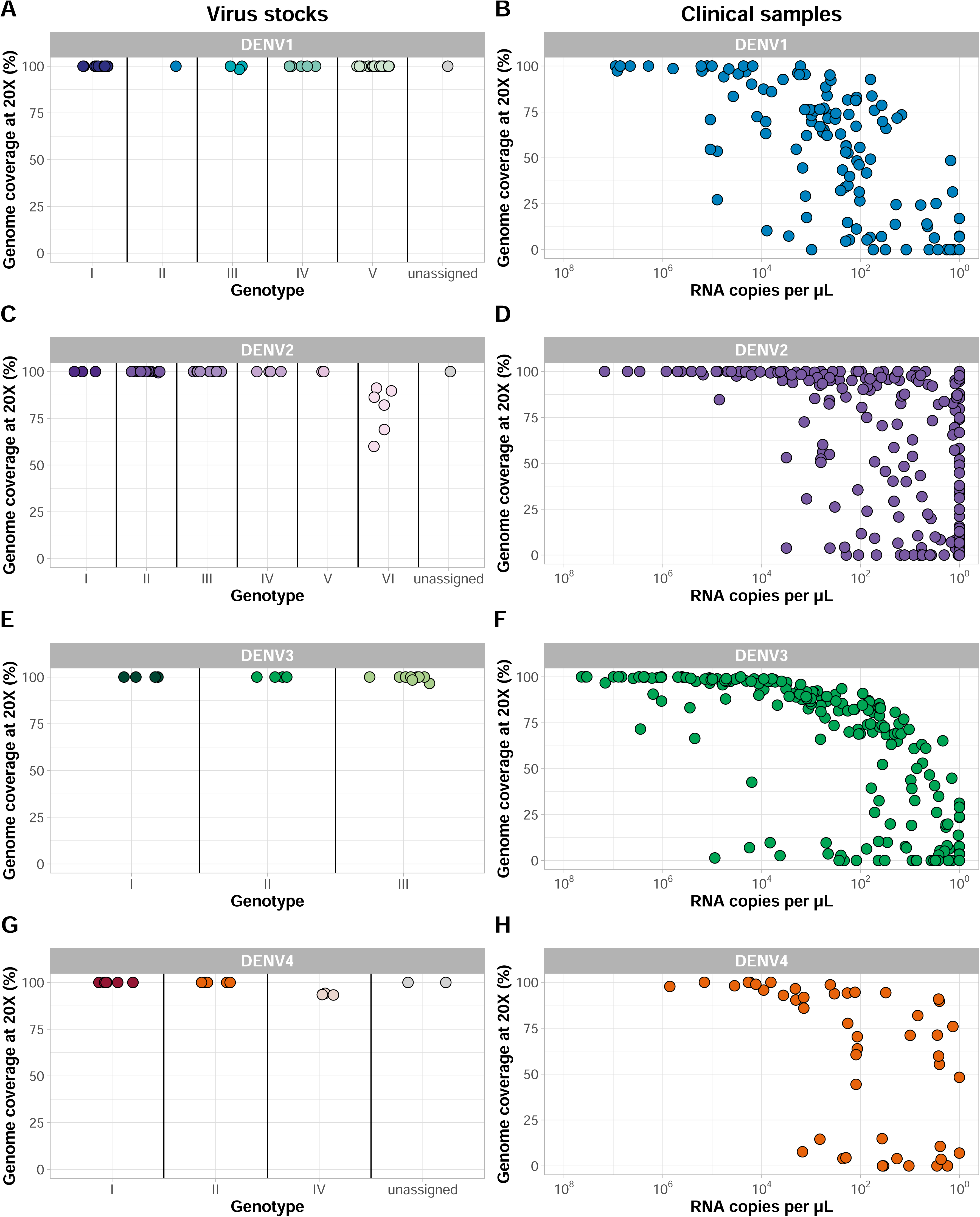
Percent genome coverage at a depth of coverage of 20x for dengue virus stocks and clinical specimens. A/C/E/G) Initial validation of the serotype-specific approach was validated by sequencing virus stocks representing the majority of genotypes within each of the four serotypes. Dengue virus 2 genotype VI and dengue virus 4 genotype IV comprise sylvatic viruses, which were the only genotypes for which genome coverage was consistently below 100%. B/D/F/H) Additional sequencing was done from clinical specimens of patients diagnosed with dengue. Specimens comprised a wider range of RNA titers, and lower genome coverage with lower RNA copies. Phylogenetic trees showing the genome sequences used for validation are shown in **Fig. S3 (virus stocks) and Fig. S4 (clinical specimens)**.

Next, we validated the DengueSeq primer schemes with clinical specimens to determine how they would work with various levels of sample quality and virus copies. We tested 600 sera samples of dengue-infected individuals from the Florida Department of Health, mostly from travelers, that also included many of the defined genotypes (**Fig. 2, Fig. S4**). Before sequencing, we determined the serotype for each sample using the CDC dengue multiplex RT-qPCR assay. We then selected the corresponding serotype-specific primer scheme for sequencing. For all four serotypes, we generated near-complete genomes for samples with high virus copies (∼10^4^ copies per μL or higher), with a clear drop-off in genome coverage with decreasing virus copies (∼10^2^ copies per μL; **Fig. 2A, C, E, G**). Importantly, we noticed that several dengue virus genomes had mismatches with the CDC dengue multiplex RT-qPCR primers and probes, resulting in decreased PCR efficiency and inaccurate measurements of virus copies. The impact of these mismatches was evident for DENV2 and DENV4, where we could often sequence viruses that were undetected by the RT-qPCR assay, and makes it difficult to set a threshold for including samples for sequencing using this assay. However, we show that our serotype-specific primer schemes can be used to successfully sequence dengue virus from clinical specimens.

### Pan-serotype primer scheme validation

The majority of clinical diagnostics used for dengue virus do not provide information on the serotype [17,18]. Thus, we developed a pan-serotype version of DengueSeq to facilitate whole-genome sequencing of clinical specimens of unknown serotypes for streamlined high-throughput sequencing. By combining the serotype-specific primer pools 1 and 2 at an increased concentration of 20 μM to account for a dilution effect, we created the two pan-serotype primer pools (**Fig. 1A**). We resequenced a subset of undiluted and diluted virus stocks as well as clinical specimens with the pan-serotype approach to compare genome coverage with the serotype-specific approach. We used a threshold of 70% genome coverage to constitute “successful sequencing”, which is a balance between obtaining as many genomes as possible while maintaining the accuracy required for correct phylogenetic placement [13]. Of the genomes with >70% coverage with the serotype-specific primer scheme (**Fig. 2**), we found that 84/90 (93.3%) of DENV1, 152/155 (98.1%) of DENV2, 37/38 (97.4%) of DENV3, and 47/47 (100%) of DENV4 met the coverage threshold with the pan-serotype approach (**Fig. 3**). When further comparing the coverage of these genomes, we found no significant differences in median genome coverage between the serotype-specific and pan-serotype approaches for either virus stocks or clinical specimens (Kruskal-Wallis tests, P>0.05 for all pairwise comparisons after Bonferroni correction; **Fig. S5**). These findings suggest that both the serotype-specific and pan-serotype approaches perform similarly when sequencing virus stocks or clinical specimens across a range of virus copies. For high-throughput and streamlined sequencing of dengue virus, we recommend using the pan-serotype approach for sequencing any dengue virus serotype.

**Fig 3:**
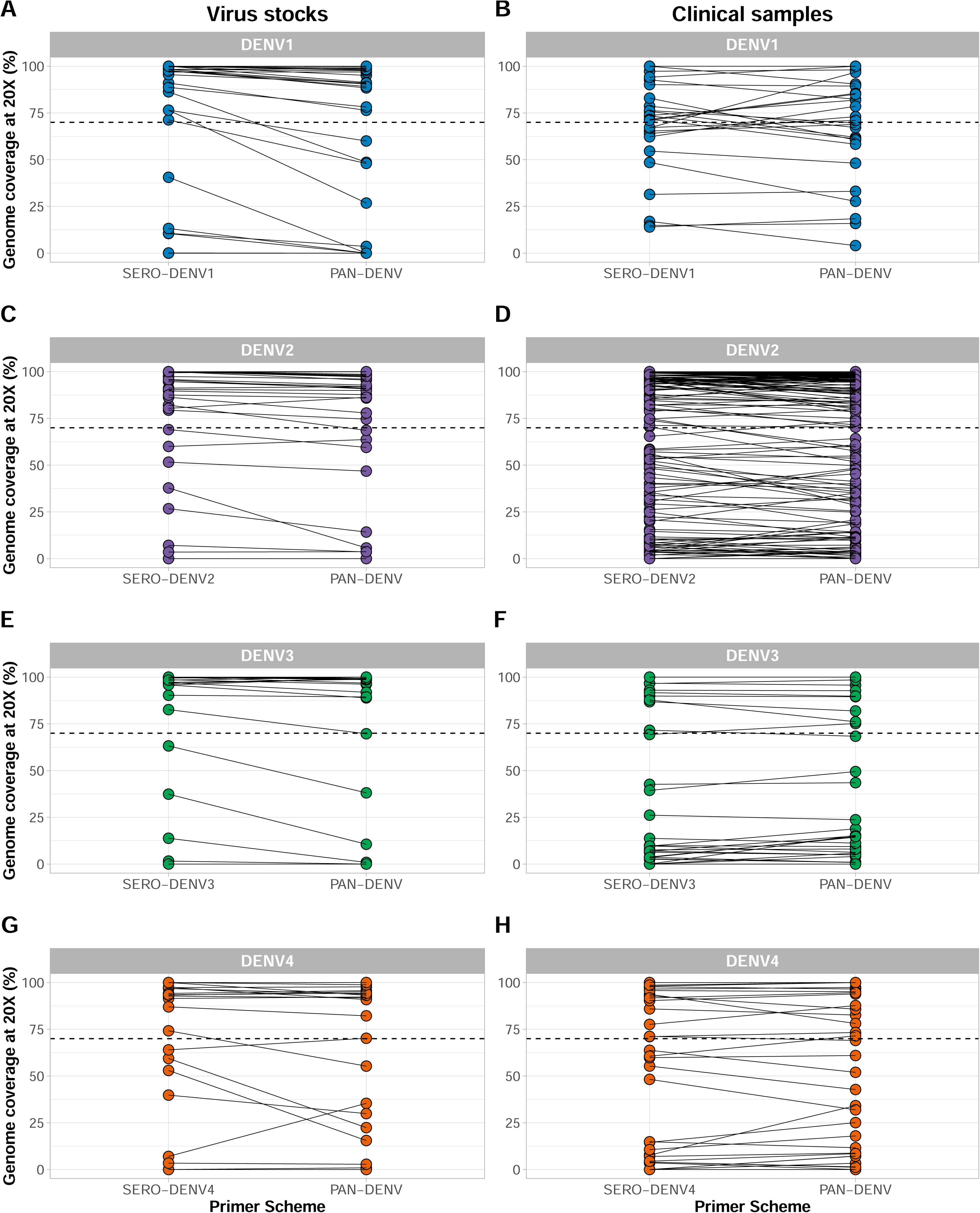
Comparison of serotype-specific and pan-serotype sequencing approaches. Dengue virus stock and clinical specimens were sequenced with both the serotype-specific and pan-serotype virus approaches to compare genome coverage between both methods. The pan-serotype approach was created by mixing the serotype-specific primer schemes into two pools (pool 1 and pool 2 for each of the combined schemes). All samples were tested with the CDC DENV1-4 multiplex qPCR assay, and libraries were prepared using the Illumina COVIDSeq test kit with serotype-specific or pan-serotype primer schemes. Connecting lines between dots indicate the same samples sequenced with both approaches and the dashed line indicates our threshold of 70% genome coverage.

### Limits of detection

We determined the limits of detection to evaluate how DengueSeq performs compared to schemes designed for other viruses. As we described above, mismatches with some of our tested viruses to the CDC dengue multiplex RT-qPCR assay led to inaccurate measurements of virus copies. To account for this, we selected viruses that do not contain impactful mismatches to determine the limits of detection for DengueSeq. For all four serotypes, we estimate that the limit of detection to achieve at least 70% genome coverage ranged between 10^1^-10^2^ RNA copies/μL (**Fig. 4A, C, E, G**). When comparing the limits of detection with other viruses, we found that the primer schemes for SARS-CoV-2 (**Fig. 4B**) and Powassan virus (**Fig. 4D**) had similar thresholds to achieve >70% genome coverage of approximately 10^2^ RNA copies/μL, when sequencing clinical or field samples. The threshold for Zika virus (**Fig. 4F**) and West Nile virus (**Fig. 4H**) based on sequencing diluted virus stocks was slightly higher at 10^3^-10^5^ RNA copies/μL. This shows that DengueSeq is at least as sensitive as other commonly used primer schemes for whole-genome sequencing of viruses of public health importance.

**Fig. 4:**
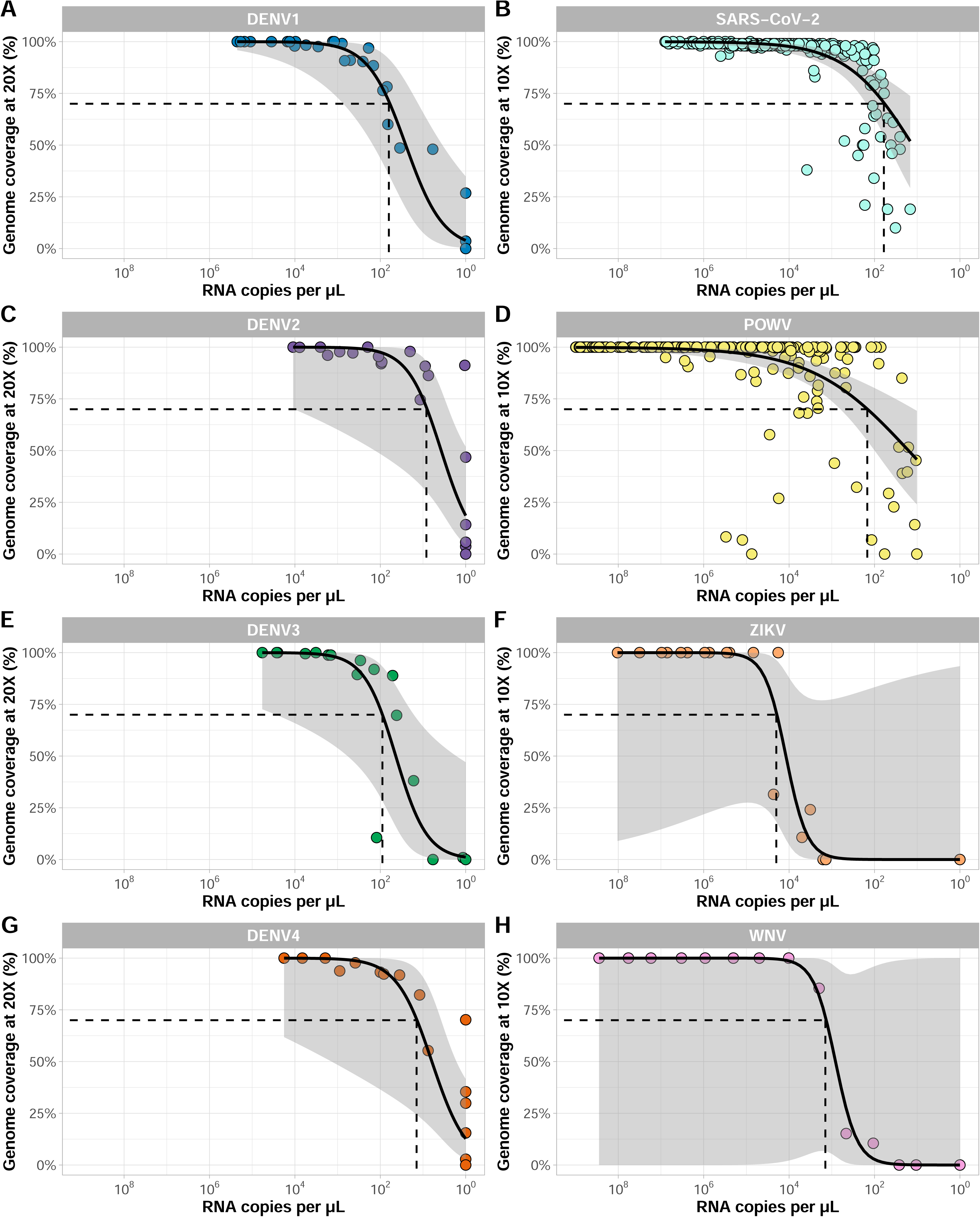
Limits of detection of pan-serotype approaches and primer schemes for other viruses. A/C/E/G) Virus stocks were diluted to represent a wide range of RNA titers and were sequenced with the pan-serotype virus approach using the Illumina CovidSeq test kit. B) Clinical specimens testing positive for SARS-CoV-2 were sequenced using the ARTIC v4.1 primer scheme using the Illumina COVIDSeq test kit. D) *Ixodes scapularis* ticks testing positive for Powassan virus were sequenced with the Powassan virus primer scheme using the Illumina COVIDSeq test kit. F) Two Zika virus stocks were sequenced with the Zika virus primer scheme using the Illumina CovidSeq test kit. H) A West Nile virus stock was sequenced with the West Nile virus primer scheme using the Illumina CovidSeq test kit. Dots represent individual samples, and a line with confidence intervals was fitted using the geom_smooth function with the ‘glm’ method from ggplot2.

### Contrived co-infections

With multiple dengue virus serotypes concurrently circulating in the same areas, there is a risk for co-infections with different serotypes [19–21]. The added advantage of using the pan-serotype primer scheme with DengueSeq is the ability to detect co-infections and sequence multiple serotypes in the same sample. When sequencing clinical specimens (**Fig. 3**), we simultaneously generated near-complete genome sequences of both DENV1 (92.9%) and DENV3 (100%) from the same specimen. To further explore the ability to sequence multiple serotypes during co-infections, we (1) created co-infections by mixing two clinical specimens containing two different serotypes and (2) mixed different concentrations of DENV1 and DENV2 stocks. With the former, we successfully sequenced two different serotypes across a range of virus copies, and the genome coverages were similar to what we generated from single infections (**Fig. 5A**). By sequencing the stock co-infections at different dilutions, we found that genome coverage was not affected by the presence of another dengue serotype across the tested range of dilutions (**Fig. 5B**). Thus, by sequencing contrived clinical and stock virus co-infections we showed that the pan-serotype DengueSeq approach has an added benefit of reliably detecting multiple serotypes in the same sample.

**Fig. 5:**
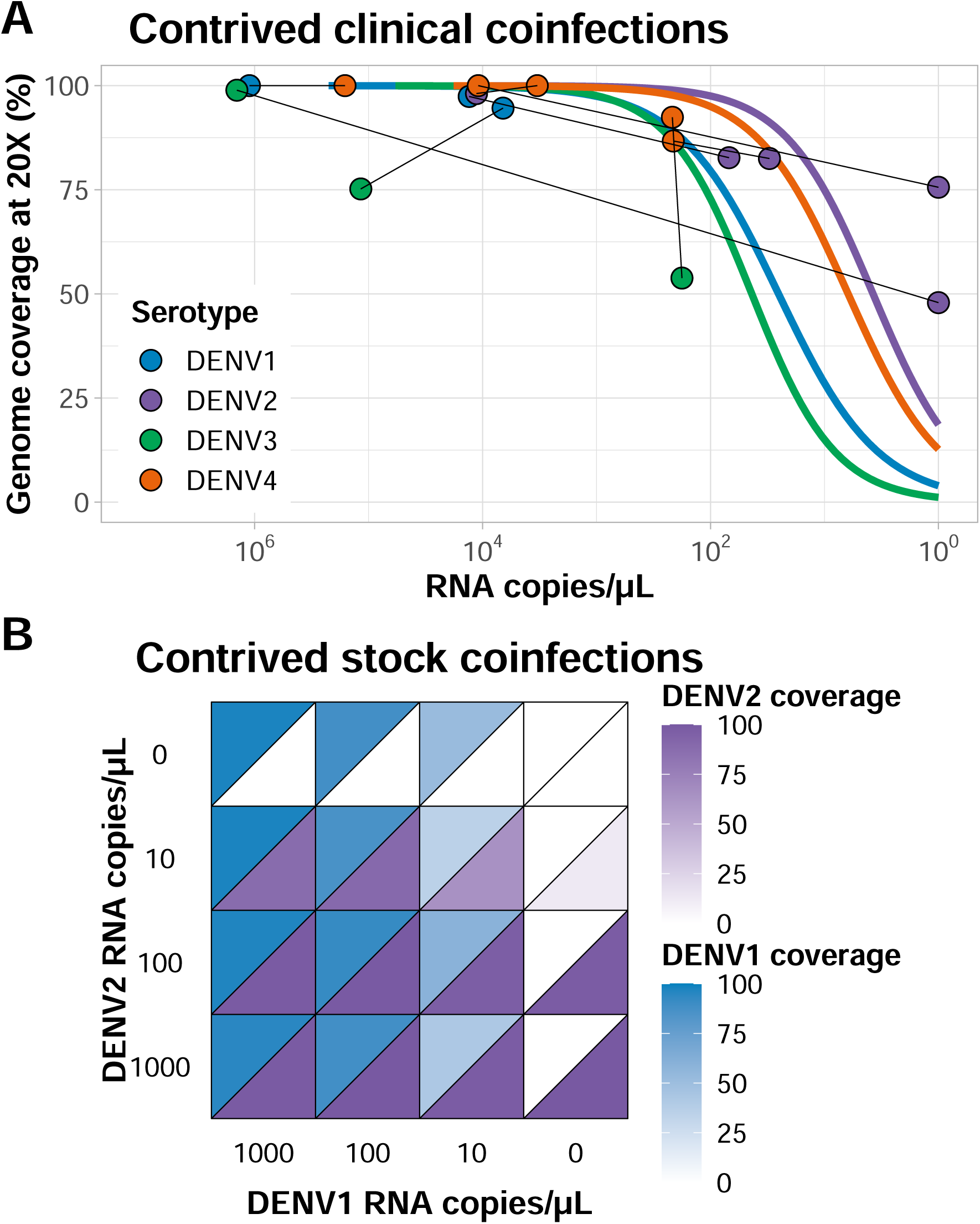
Detection of dengue virus coinfections using the pan-serotype primer scheme. A) Clinical specimens containing different dengue virus serotypes were mixed to create contrived coinfections. A total of 8 contrived samples were created by mixing two different clinical specimens. Dots represent individual samples connected with lines to indicate mixed pairs and colored lines are expected genome coverage estimated from fitted logistic regression lines determined based on coverage data from Fig. 4. B) Two virus stocks (serotypes 1 and 2) were serially diluted and mixed at different concentrations to represent a range of virus titers in contrived stock coinfections.

### Other library prep kits

Previous studies have shown that amplicon primer schemes can be used with a variety of library prep kits [8,9,22]. Here, we validated DengueSeq primarily with the Illumina COVIDSeq test kit because it enables high-throughput sequencing without manual normalization steps, and many labs around the world have implemented this kit during the COVID-19 pandemic. However, as some labs prefer different amplicon sequencing workflows, we tested DengueSeq with different library prep kits and sequencing instruments. We validated the pan-serotype primer scheme with three additional workflows in different labs: (1) Illumina Nextera XT library prep kit and the Illumina MiSeq at the Florida Department of Health, (2) New England Biolabs NEBNext and the Illumina NovaSeq at the Yale School of Public Health, and (3) “Midnight” protocol using the Oxford Nanopore Technologies rapid barcoding kit and the GridION at the Amsterdam University Medical Centers (**Fig. 6**). Although we cannot make direct comparisons, we showed that the pan-serotype primers can be used across a variety of amplicon sequencing workflows and in different labs. Some of the samples that were sequenced with the NEBNext library prep kit had lower-than-expected genome coverage. An imbalance in the final concentrations of individual libraries in the final pool due to manual normalization may explain the higher variation observed and can be further optimized. Testing the pan-serotype primer scheme across library prep kits, sequencing instruments, and in different labs shows that DengueSeq can be integrated into existing amplicon sequencing workflows with minimal change to the overall protocol.

**Fig. 6:**
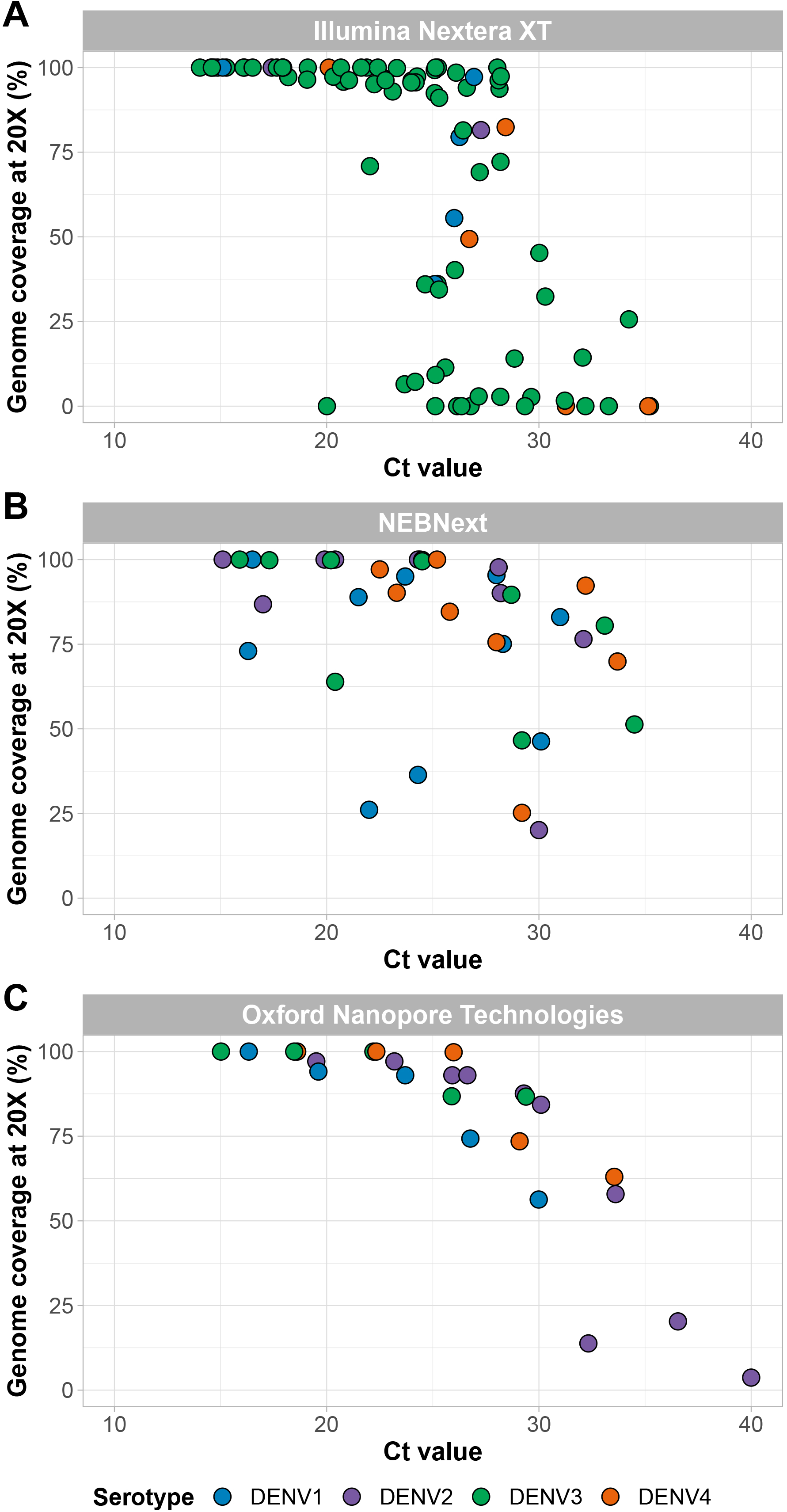
Alternative library prep methods used with the pan-serotype primer scheme. A) The Florida Department of Health sequenced clinical specimens using the pan-serotype primer scheme with the Illumina Nextera XT kit. B) Clinical specimens were sequenced using the pan-serotype primer scheme with the NEB NEBNext library prep kit for Illumina. C) Amsterdam UMC sequenced virus stocks using the pan-serotype primer scheme with the “Midnight” protocol and sequenced on the Oxford Nanopore Technologies GridION.

## Discussion

Expanded genomic surveillance of dengue virus is critically needed to gain insight into currently circulating genetic diversity, emergence and spread of new lineages, and to evaluate the effectiveness of newly implemented control tools. We developed DengueSeq, a whole-genome pan-serotype sequencing protocol, and associated bioinformatics pipeline to facilitate expanded genomic surveillance of dengue virus by utilizing widely implemented amplicon sequencing workflows. We designed the multiplexed primers to amplify viruses comprising the known diversity of dengue virus causing human outbreaks (**Fig. 1**), and then validated the approach with a panel of genetically diverse virus stocks and clinical specimens from all four serotypes (**Fig. 2**). We showed that DengueSeq can be used with any of the four serotype-specific primers or as a combined pan-serotype approach (**Fig. 3**). We estimate that 10^1^-10^2^ virus copies per μL is required to generate at least 70% genome coverage for all four serotypes, which is similar to the ARTIC SARS-CoV-2 assay (**Fig. 4**). Furthermore, we demonstrate that the pan-serotype protocol can be used for samples with unknown serotypes and is able to sequence multiple serotypes in the same sample (e.g. coinfections; **Fig. 5**). DengueSeq provides a unified approach for whole genome amplicon sequencing of dengue virus that can be used with a variety of library prep methods and sequencing instruments, with minimal change needed to currently established amplicon sequencing workflows (**Fig. 6**). The ability to combine the four serotype-specific primer schemes into a unified pan-serotype approach demonstrates the proof of principle that amplicon sequencing schemes can be scaled to broad-spectrum virus sequencing workflows.

The development of PrimalScheme has enabled researchers to develop custom primer schemes for amplicon-based sequencing. DengueSeq is not the first nor the only primer scheme available for dengue virus, and several other custom dengue virus primer schemes have recently been developed [23–28]. What makes DengueSeq different from other custom primer schemes is that we (1) designed our primer schemes to include global genetic diversity within each of the serotypes, (2) performed comprehensive validation with samples representing genetic diversity within each serotype, different sample types, and a range of virus copies, and (3) validated a pan-serotype approach that can be used to perform whole genome sequencing of all four serotypes without prior serotyping. Additionally, several commercial virus target enrichment panels are currently available for broad virus surveillance. The main advantage of these panels is that they capture a large number of viruses, but the disadvantages are the relatively high cost and lower sensitivity as compared to amplicon sequencing. Thus, several solutions are available that can potentially be used for dengue virus sequencing. DengueSeq is particularly useful for labs that have already implemented amplicon sequencing workflows and/or are looking for a relatively low-cost, sensitive, and high-throughput sequencing system to conduct genomic surveillance of dengue virus from clinical samples that typically have low virus copies. These features of the DengueSeq approach would also make it easily deployable for dengue genomic surveillance, especially in low-income settings with scant genomic data.

High dengue virus genetic diversity poses challenges to successful whole genome sequencing. Our approach of combining the four serotype-specific primer schemes into the pan-serotype approach allows us to capture the current genetic diversity present within distinct genotypes in the four serotypes. However, we cannot predict whether the current primer schemes will capture newly emerging lineages. This scenario is exemplified by the lower coverage of sylvatic genomes (**Fig. 2C, 2G**), which enables identification but not whole genome sequencing. However, updating the DengueSeq scheme to include newly characterized lineages can improve the sequencing coverage of these emerging lineages. Several other factors may also affect the future performance of DengueSeq. To account for the high genetic diversity of dengue virus, we initially designed “degenerate” primer schemes that included ambiguous nucleotides at divergent primer-binding positions. The inclusion of ambiguous nucleotides did not improve the sensitivity of the primer schemes, and, therefore, we proceeded with validation of the non-degenerate (original) primer schemes. We determined the limits of detection ranging between 10^1^-10^2^ RNA copies/μL to achieve at least 70% genome coverage. However, we should interpret these thresholds carefully as the threshold for DengueSeq when sequencing clinical specimens may need to be more conservative depending on sample quality, specimen type, available resources, etc. We recommend that laboratories quantify the relationship between genome coverage and RNA copies to determine their threshold, as this would allow them to prioritize specimens for sequencing. The choice of reference genomes for reference-based alignment of sequencing reads had a significant impact on genome coverage of highly divergent genomes. By using DENV2 and DENV4 sylvatic references in the bioinformatic analysis, we generated higher genome coverage than when aligning reads to the non-sylvatic reference genomes. Future development of virus-agnostic bioinformatics pipelines from amplicon-based sequencing would be beneficial to ensure proper alignment of divergent sequencing reads. The choice of library prep methods is another consideration for labs wanting to implement DengueSeq. We showed that the pan-serotype approach can be used with a variety of library prep methods for sequencing on Illumina and Oxford Nanopore Technologies instruments. For some of the methods that require manual normalization and pooling, we found higher variation in genome coverage, which is likely the result of an imbalance in the individual libraries that were pooled. We recommend that laboratories use DengueSeq to expand currently-implemented amplicon sequencing workflows, e.g., for SARS-CoV-2, to include dengue virus.

## Conclusions

We demonstrated that DengueSeq is a unified, sensitive, and high-throughput whole genome amplicon sequencing protocol for all four dengue virus serotypes. We designed and validated DengueSeq with a broad panel of samples to represent the breadth of genetic diversity, different sample types, and a range of virus copies. Our findings show that DengueSeq can be used with currently established amplicon sequencing workflows and provide a proof of principle that multiple primer schemes can be combined into unified approaches to improve the sequencing of multiple dengue serotypes. The use of DengueSeq will facilitate genomic surveillance of dengue virus, which is critically needed to understand the increasing scale and frequency of outbreaks and the effectiveness of control measures.

## Methods

### Primer scheme design

Four separate serotype-specific primer schemes were designed using PrimalScheme. For each serotype, genetically diverse genomes were selected to represent the distinct genotypes **(Fig 1B)**. Genotypes with only incomplete genomes available and highly divergent genotypes were excluded (e.g. DENV1 genotype II, DENV2 genotype IV and VI (sylvatic), and DENV4 genotypes III and IV (sylvatic)), until a primer scheme with high coverage across the genome was found. Each primer scheme consists of 35-37 primer pairs with an average amplicon length of 400 bp. The primer schemes can be used as a serotype-specific approach when the serotype is known (working concentration of 10 μM), or combined at equal concentrations as a pan-serotype approach (working concentration of 20 μM). Primer sequences, reference files, and bed files are available on protocols.io [29].

### Virus stocks for validation

Genetically diverse virus stocks with representatives for all dengue virus serotypes and genotypes from the Yale Arbovirus Research Unit and World Reference Center for Emerging Viruses and Arboviruses collections were kindly shared by the Connecticut Agricultural Experiment Station and the University of Texas Medical Branch. The total collection of virus stocks used for validation consisted of 46 DENV1, 46 DENV2, 17 DENV3, and 18 DENV4 isolates. Viruses were sequenced with the serotype-specific and pan-serotype approaches. The dengue multiplex RT-qPCR assay as developed by the CDC was used to determine serotype and PCR Ct values for each stock [15]. Virus stocks were serially diluted to represent a range of Ct values. Serial ten-fold dilutions of quantified synthetic dengue virus 1-4 controls obtained from ATCC were used to convert Ct values to RNA copies per μL.

### Clinical specimens for validation

De-identified remnant clinical specimens were provided by the Florida Department of Health and nucleic acid was extracted by Florida Gulf Coast University. Clinical specimens were collected between 2010-2023 and comprised locally acquired cases as well as travel-related cases. Nucleic acid was extracted using the Qiagen QIAamp viral RNA mini kit. The dengue multiplex RT-qPCR assay, as developed by the CDC, was used to determine serotype and PCR Ct values for each specimen [15]. The total collection of clinical specimens used for validation consisted of 121 DENV1, 214 DENV2, 219 DENV3, and 46 DENV4 samples.The serotype-specific approach was used to sequence all specimens, and a smaller subset was re-sequenced using the pan-serotype approach, as described below.

### Amplicon sequencing

Initial amplicon sequencing validation was performed at the Yale School of Public Health. To test the primer scheme in diverse lab settings and with different library prep kits, additional testing was performed by the Florida Department of Health and the Amsterdam University Medical Centers. A detailed protocol is available on protocols.io [29].

At the Yale School of Public Health, initial validation of the primer schemes was done with (diluted) virus stocks and de-identified remnant clinical specimens. Samples were sequenced using the Illumina COVIDSeq test (RUO version) by replacing primers as part of the kit for the serotype-specific or pan-serotype primer pools. A negative template control was included during cDNA synthesis and amplicon generation for each sequencing run. Pooled libraries were sequenced on the Illumina NovaSeq 6000 (paired-end 150) at the Yale Center for Genome Analysis, targeting 1 million reads per sample. Consensus genomes were generated using the newly developed bioinformatics pipeline described below. Sequencing data and consensus genomes are available in BioProject PRJNA951702 and PRJNA1001374.

At the Florida Department of Health, validation of the primer schemes was done with 84 clinical specimens consisting of serum from individuals diagnosed with dengue. Nucleic acid was extracted from clinical specimens using the Qiagen Qiacube DSP Viral RNA Mini and tested with the CDC Dengue Virus Typing (DENV-1/2/3/4) FDA IVD assay on the Thermo Fisher ABI 7500. Libraries were prepared for sequencing using the Illumina Nextera XT library prep kit, and sequenced on the Illumina Miseq instrument (251x2), targeting 250,000 reads (range of 150,000-400,000) per sample. Consensus genomes were generated with the newly developed bioinformatics pipeline. Sequencing data and consensus genomes are available in BioProject PRJNA973096.

At Amsterdam University Medical Centers, the primer scheme was validated using five cultured dengue strains passaged on VeroE6 cells. Nucleic acids were extracted from 200 μL of culture supernatant using the Qiagen QIAamp viral RNA mini kit with an elution volume of 50 μL. Viral loads were determined using the Roche LightMix modular dengue virus kit on the Roche Lightcycler 480, according to the manufacturer’s protocol. Sequencing libraries were prepared for sequencing using the Oxford Nanopore Technologies (ONT) rapid barcoding kit 96 following the “midnight” protocol [30], and sequenced on the Oxford Nanopore Technologies GridION, using an R.9.4.1 flowcell, targeting 30,000 reads per sample. Consensus genomes were generated at a depth of 20x with the ARTIC bioinformatics pipeline [31], by using the dengue virus 1-4 reference genomes (dengue virus 1: NC_001477, dengue virus 2: NC_001474, dengue virus 3: NC_001475, dengue virus 4: NC_002640) and corresponding BED files.

### Bioinformatics pipeline

Raw demultiplexed read data generated from sequencing amplicons were processed using a pip-installable, Snakemake-based pipeline to generate consensus genome sequences of the dengue serotypes in the clinical sample (**Fig. S1**). The bioinformatics pipeline used for this analysis is available on Github (https://github.com/grubaughlab/DENV_pipeline/tree/v1.0), and is based on Python v3.9 and bash scripts. The input of the pipeline are the forward and reverse FASTQ read files for each sample, and a set of BED and FASTA files for the primer positions and corresponding reference sequences. The default for the pipeline for the latter is the reference sequences and primers for DENV1 (GenBank accession: NC_001477.1), DENV2 (GenBank accession: NC_001474.2), DENV3 (NC_001475.2), DENV4 (GenBank accession: NC_002640.1), DENV2 sylvatic (GenBank accession: EF105380.1) and DENV4 sylvatic (GenBank accession: JF262780.1) for the protocol described in this paper. The reference sequences are selected to represent the diversity of all known dengue serotypes so that the best match can be found with the virus in the clinical sample.

Each set of reads is mapped against each reference sequence separately (non-competitive alignment) using BWA v0.7.17-r1188 [32], and primers corresponding to each reference are trimmed using iVar v.1.4 [9]. If no reads map successfully, then the rest of the algorithm is skipped. BAM files are then sorted and indexed using samtools v1.17 [33], and the consensus nucleotide sequence is generated using samtools and iVar. A frequency threshold of 0.75 for the single nucleotide variants included in the consensus sequence and a per-base depth of at least 10 mapped reads is used by default, although both can be changed by the user. We investigated these two parameters and found that these values had the best balance of accuracy and reliability.

Once the consensus genome sequence is generated, it is aligned to the same reference sequence using Nextalign v2.1 [34]. If there are too few reads, Nextalign may fail to generate the alignment; MAFFT v7.520 is used in these circumstances [35]. The genome coverage or genome completeness, i.e., the percentage of bases in the aligned sequence not containing ambiguous bases, is then calculated using a custom Python script utilizing the BioPython module [36]. Finally, variants and per-base depth at each position in the generated mapping output (sorted BAM file) are identified using samtools, iVar and BEDTools v2.31.0 [37].

Outputs of the pipeline include three summary tab-separated text files containing information on coverage against each possible reference sequence: one with all information, one with any “called” serotype (defined as above 50% coverage), and one with whatever the reference with the highest coverage is. Alignments in FASTA format, BAM files, and consensus sequences of called serotypes are also available, as are text files containing depth at each position of each sample for their called serotype. Finally, text files containing information on nucleotide variants against called serotypes are output.

## Data analysis and visualizations

All data analysis and plotting was performed using R statistical software v4.3.1 [38] using the ggplot2 v3.4.2 [39], dplyr v1.1.2 [40], tidyverse v2.0.0 [41], cowplot v1.1.1 [42], and scales v1.2.1 packages [43]. Differences in coverage between the serotype-specific and pan-serotype approaches were tested with Kruskal-Wallis tests with Bonferroni correction for multiple comparisons. For coverage plots, lines with confidence intervals were fitted using the geom_smooth function with the ’glm’ method from ggplot2.

For the phylogenetic trees displayed in figures 1, S3, and S4, we began by collating a background set of sequences for each serotype, which were the same background set used for primer selection. We combined these with each set of sequences of interest - i.e., the primers, the virus stocks, and the clinical sequences - using BioPython in a custom script [36], aligned them using MAFFT v7.490 [35], and manually curated the resulting alignment. We then inferred a Maximum Likelihood tree using IQTree [44] with the HKY substitution model [45]. Finally, we visualized these using the Python package baltic [46].

## Data Availability

All genomic data are available in the manuscript, supporting information, GitHub (https://github.com/grubaughlab/DENV_pipeline/tree/v1.0), and BioProjects PRJNA951702, PRJNA1001374, and PRJNA973096.

## Declarations

### Ethics approval and consent to participate

The Florida Department of Health shared de-identified remnant clinical specimens that tested positive for dengue virus for sequencing at the Yale School of Public Health. The Institutional Review Boards (IRB) from the Yale University Human Research Protection Program, Florida Gulf Coast University, and Florida Department of Health determined that pathogen genomic sequencing of de-identified remnant diagnostic samples as conducted in this study is not research involving human subjects (Yale IRB Protocol ID: 2000033281 and 2000028599).

### Consent for publication

Not applicable

## Authors’ contributions

CBFV, VH, and NDG conceptualized and designed the study; CBFV, MIB, LP, AS, ET-S, IMO, MEP, DD, MJ, MRAW, TL, SB, NC, RJ, CP, IS, JV, RZ, EK, LH, AM, and SM contributed to sample collection and data generation; CBFV, VH, CC, YD, NT, OT, and SS, analyzed the data; CBFV, VH, and NDG wrote the manuscript; all authors reviewed and approved the final manuscript.

## Acknowledgements

We thank S. Weaver, J. Plante, and K. Plante from the University of Texas Medical Branch for sharing dengue virus stocks from the World Reference Center for Emerging Viruses and Arboviruses; D. Brackney, A. Bransfield, and P. Armstrong from the Connecticut Agricultural Experiment Station for sharing dengue virus stocks from the Yale Arbovirus Research Unit Collection; New England Biolabs for providing reagents; A. Porzucek for help with proofreading the protocols.io; and P. Jack and S. Taylor for technical advice.

## Supplementary Figures

**Fig S1:**
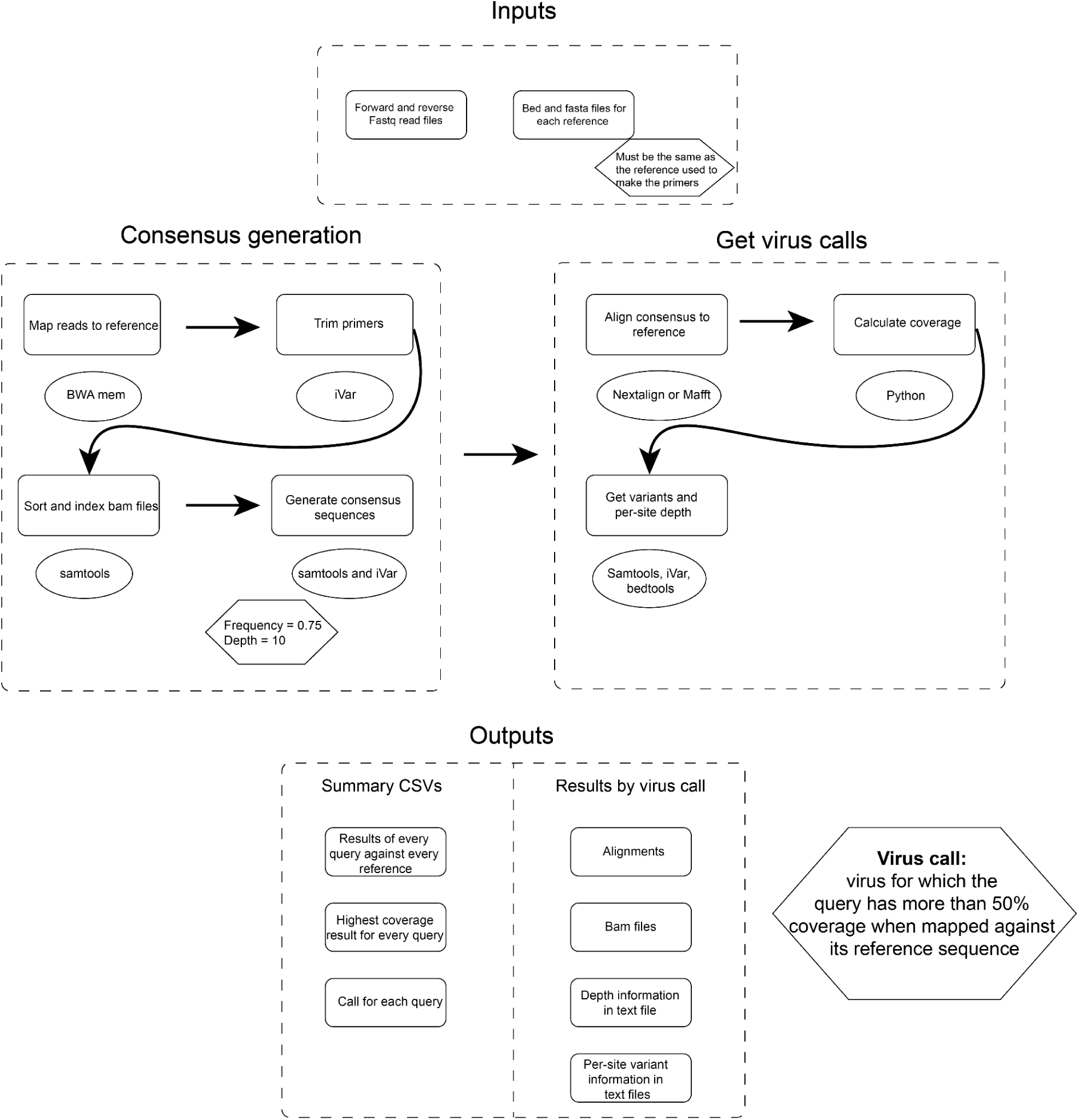
Flowchart bioinformatics pipeline. Overview of bioinformatics pipeline consisting of inputs, consensus generation, virus calls, and outputs. The pipeline is available on https://github.com/grubaughlab/DENV_pipeline/tree/v1.0.

**Fig S2:**
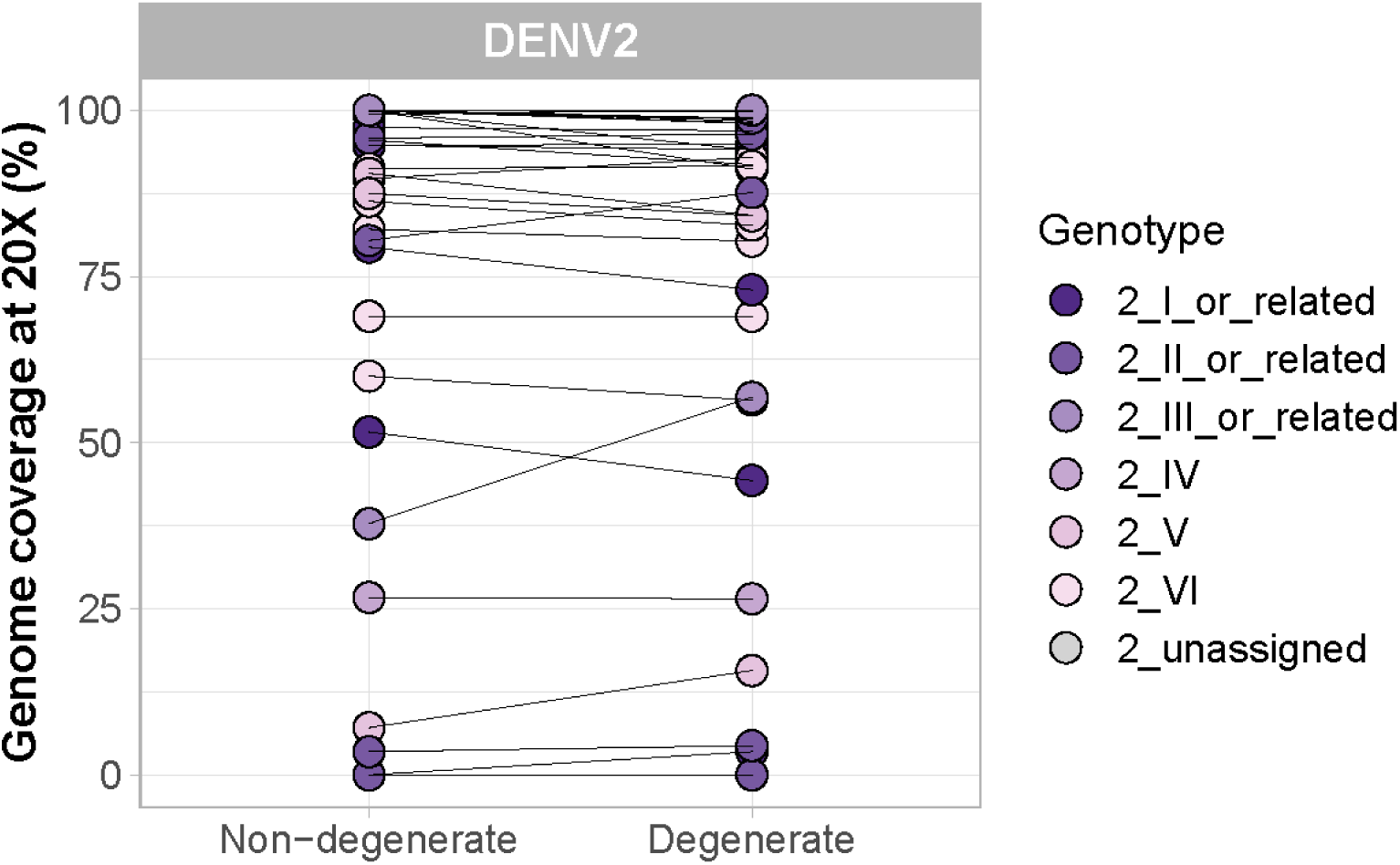
Comparison of a non-degenerate (original) and degenerate dengue virus 2 primer scheme. Pairwise comparison between dengue virus 2 stocks sequenced with non-degenerate and degenerate primer schemes. Connecting lines between dots indicate the same samples sequenced with both approaches.

**Fig S3:**
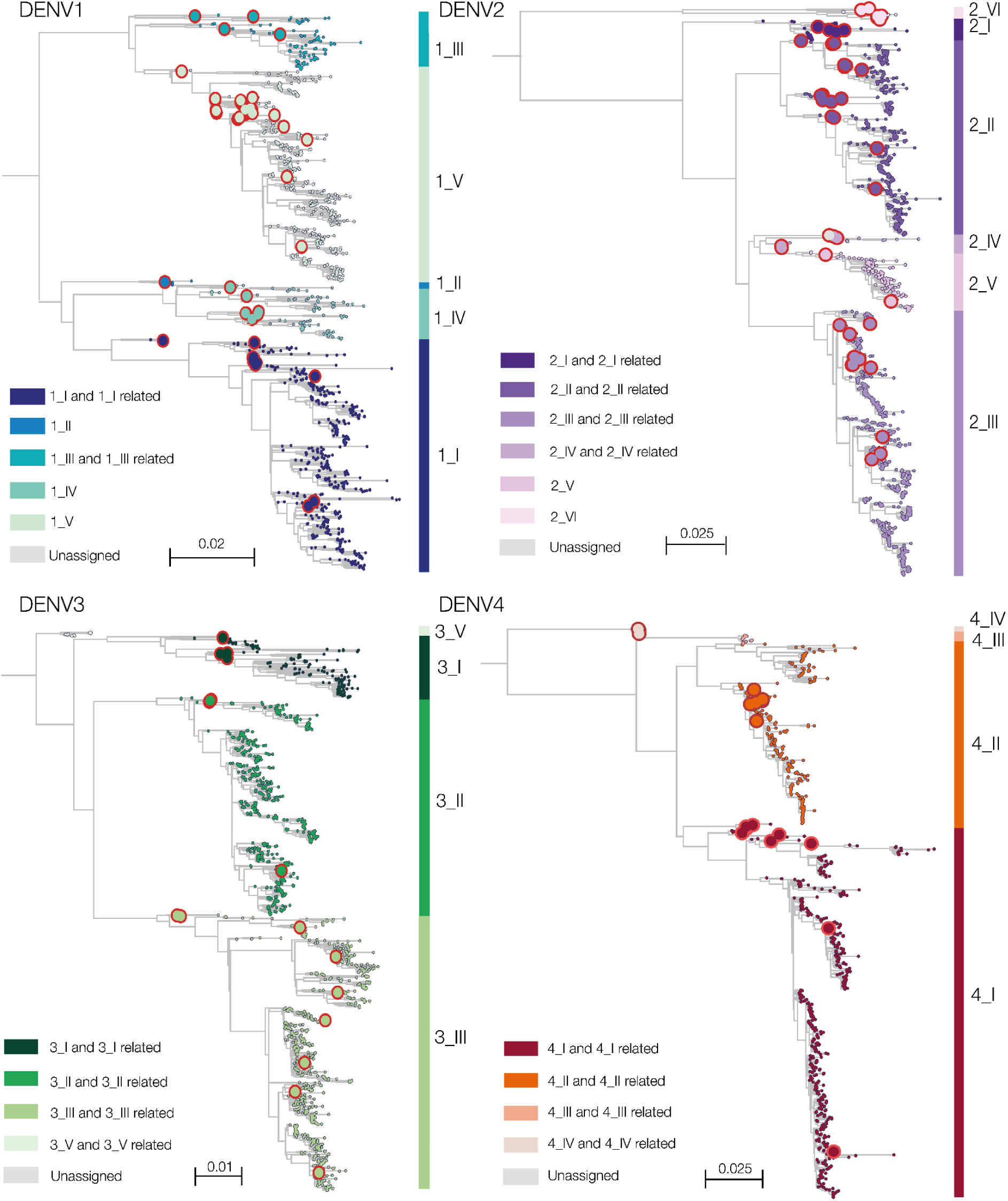
Phylogenetic tree with virus stocks used for validation. Dengue virus stocks representing genetic diversity within each of the four serotypes were used to validate DengueSeq. Highlighted in red circles are genomes that were used for validation.

**Fig S4:**
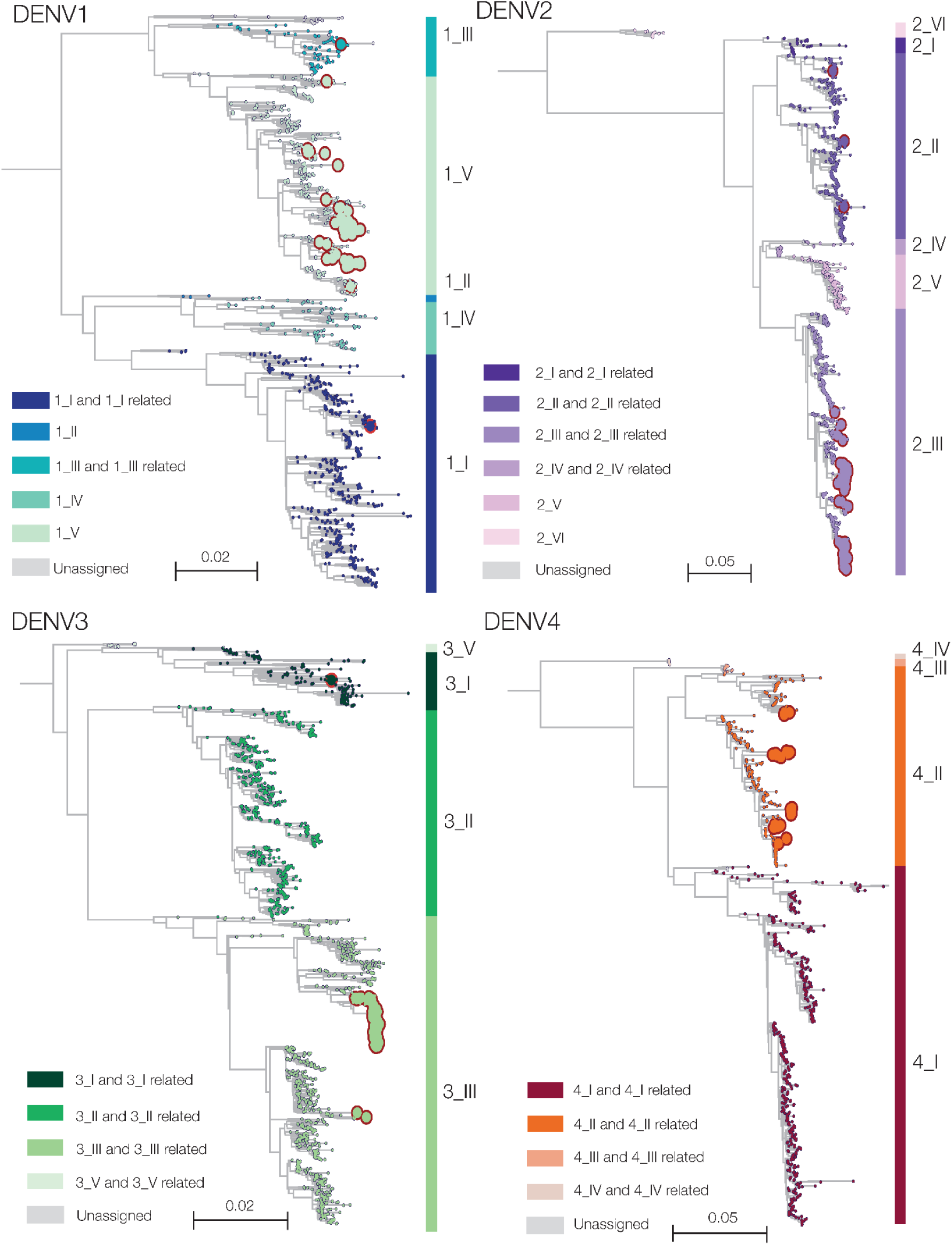
Phylogenetic tree with clinical specimens used for validation. The Florida Department of Health shared serum from dengue-positive individuals, which were sequenced with the serotype-specific and pan-serotype approaches. Highlighted in red circles are genomes with at least 70% coverage, which were used for validation.

**Fig S5:**
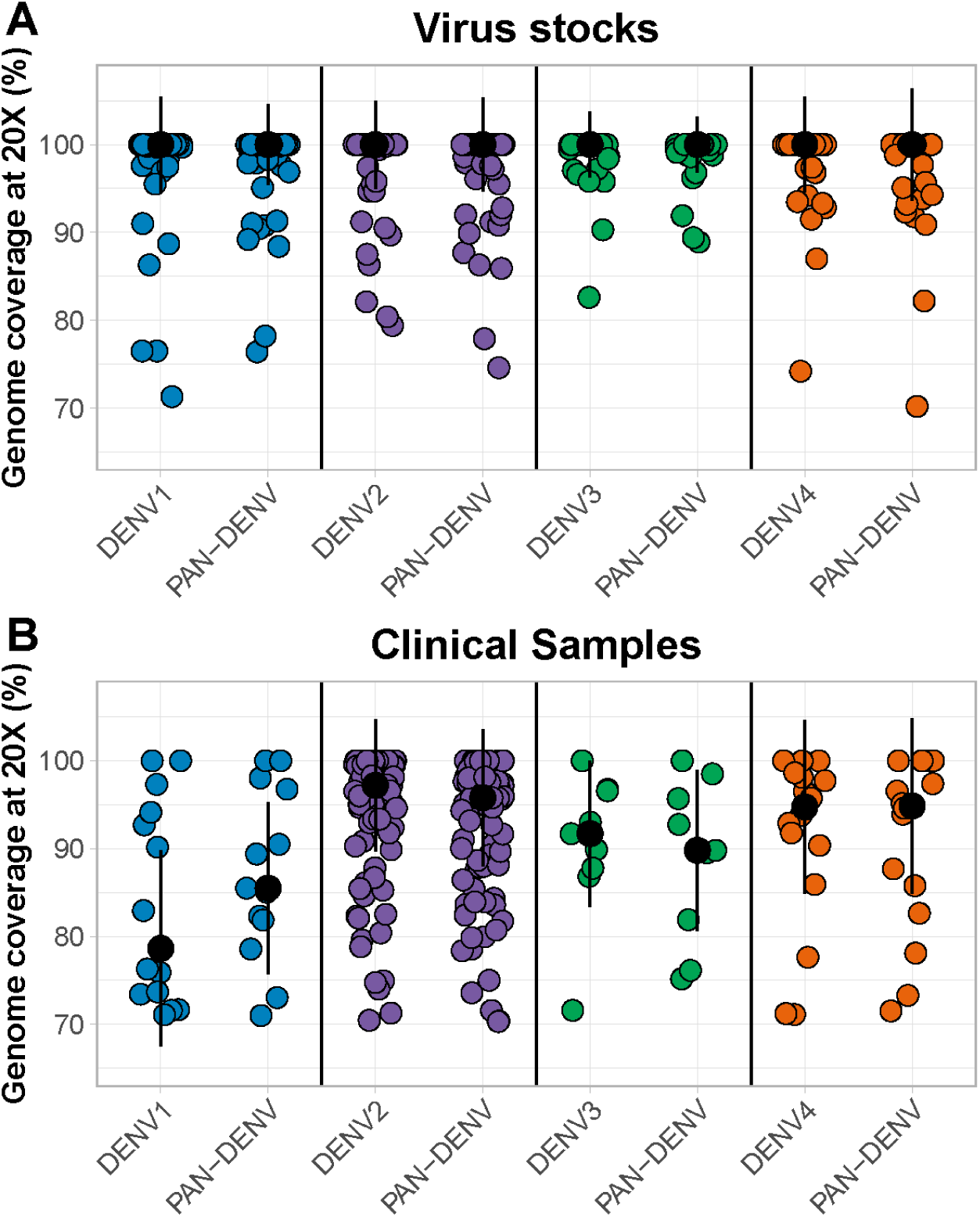
Comparison of serotype-specific and pan-serotype sequencing approaches. Genome coverage was compared between virus stocks and clinical specimens sequenced with the serotype-specific and pan-serotype approaches. Differences in coverage between the two approaches were tested with Kruskal-Wallis tests, and no significant differences were found for any of the serotypes (P>0.05 for all pairwise comparisons, after Bonferroni correction). Only genomes with coverage above the 70% threshold were included. Colored dots represent coverage for individual samples, black dots represent the median coverage, and error bars show the standard deviation.

