## Supplementary figures and images for "DengueSeq: A pan-serotype whole genome amplicon sequencing protocol for dengue virus"

### Fig S1

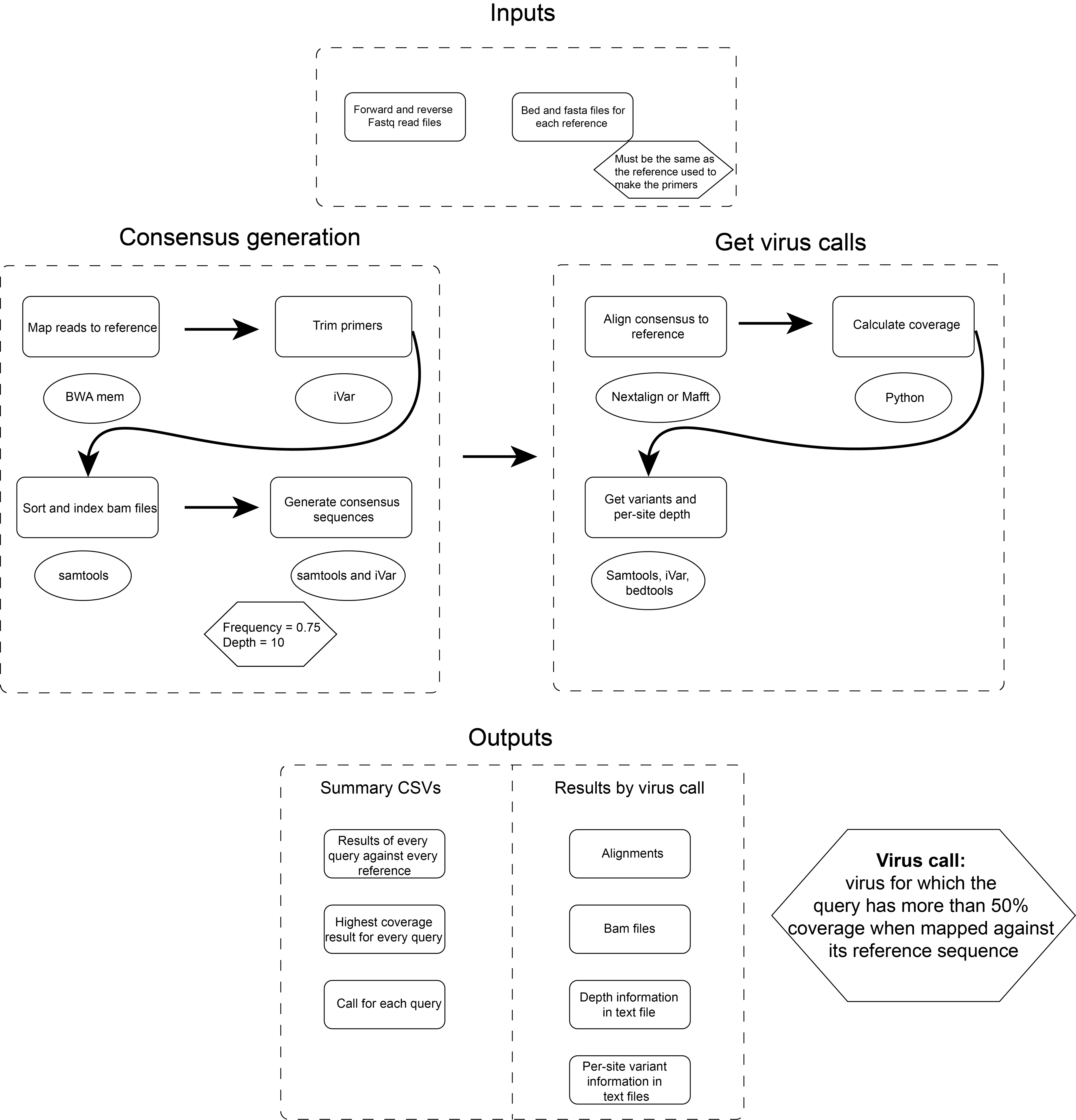

### Fig S2

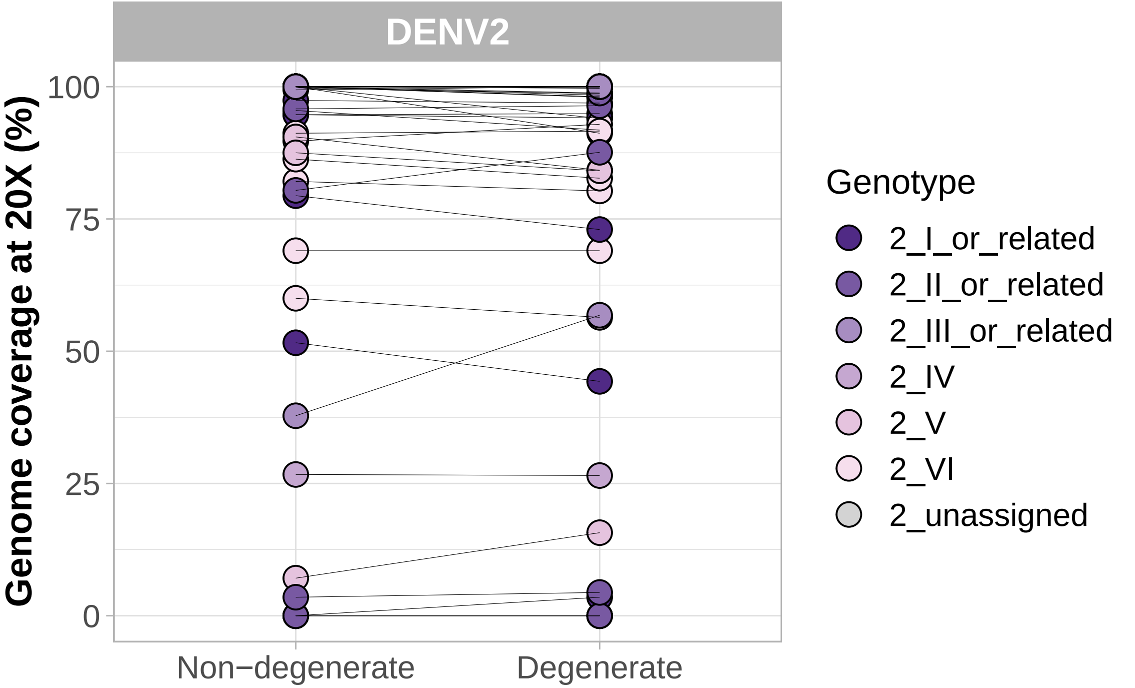

### Fig S3

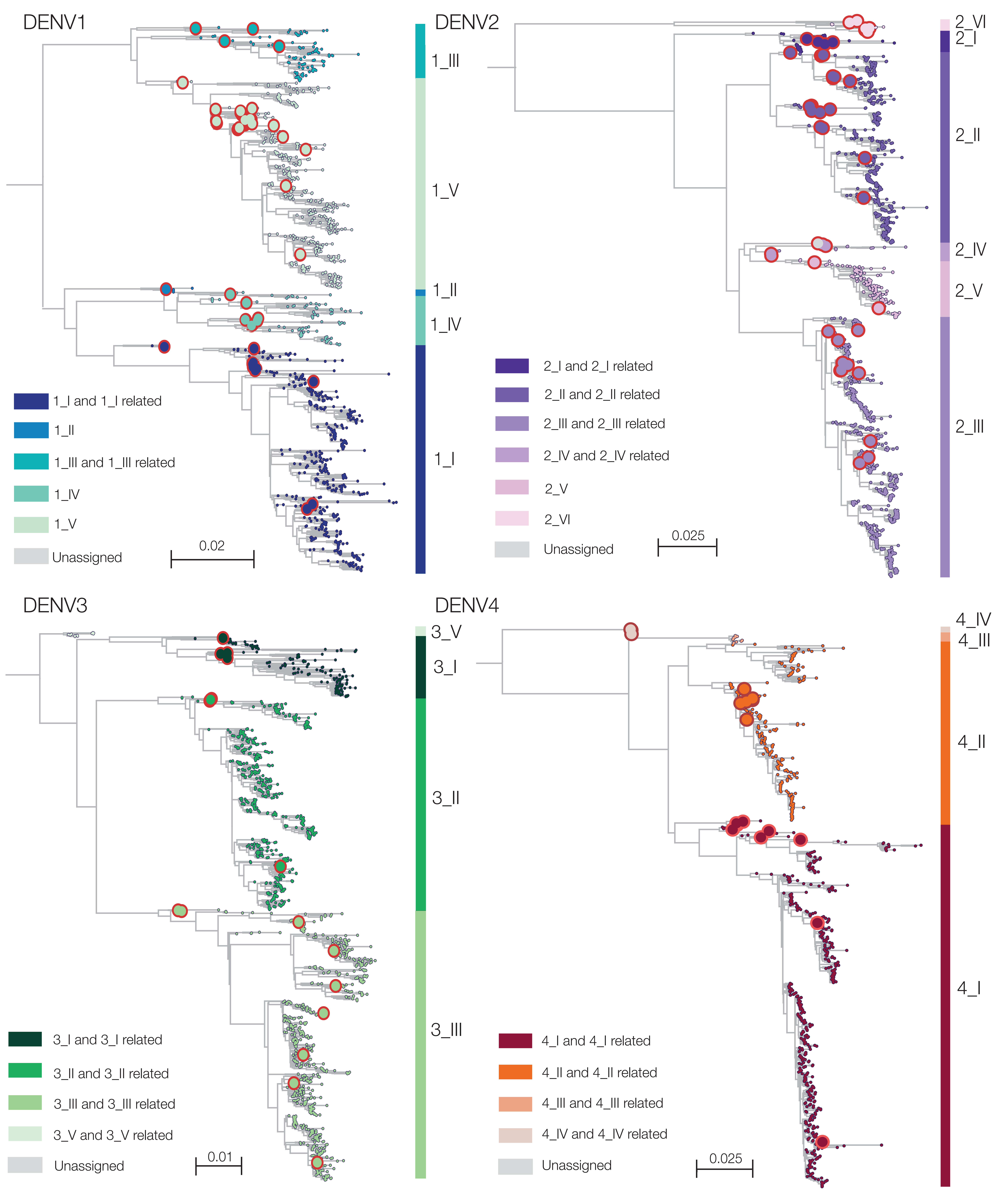

### Fig S4

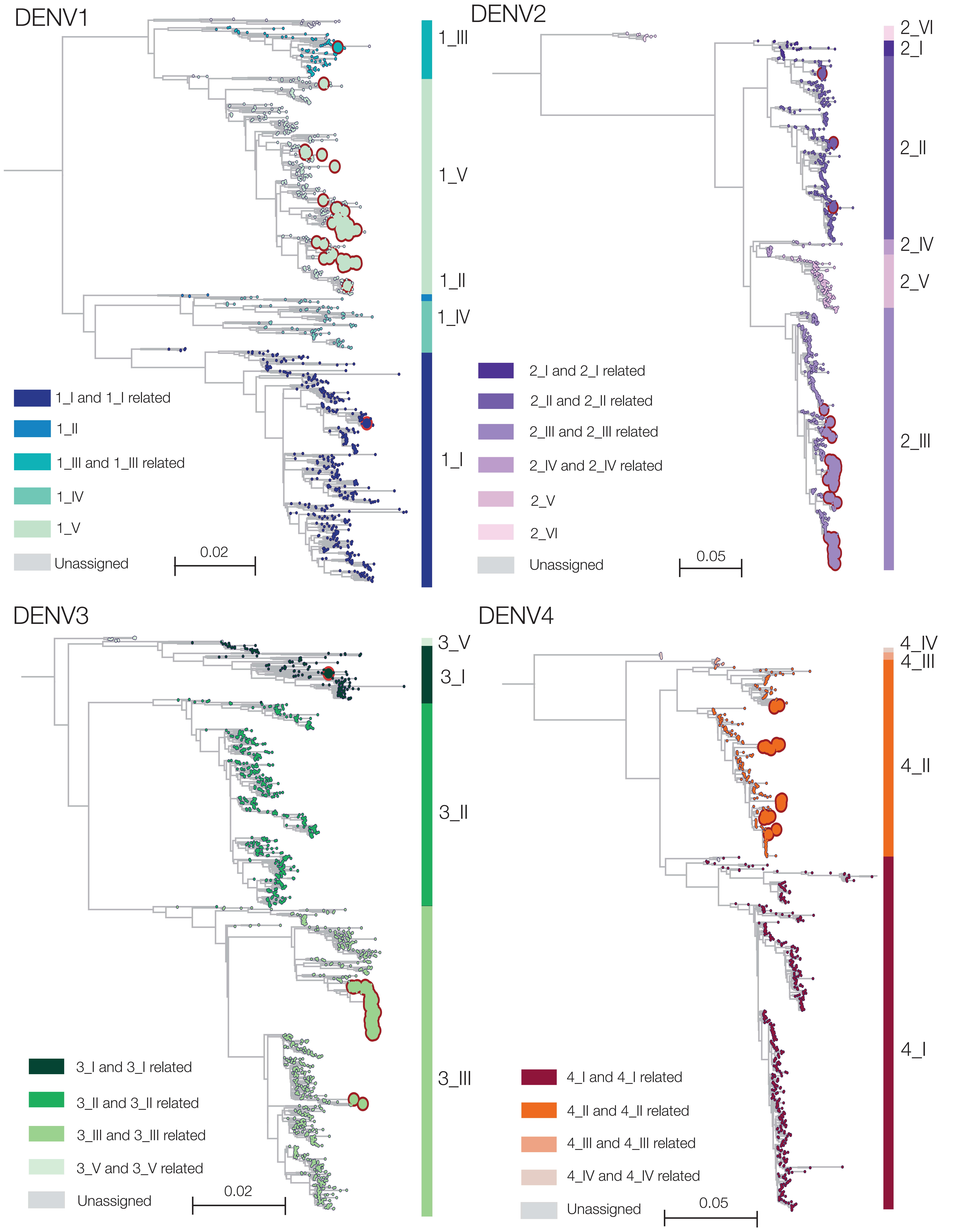

### Fig S5

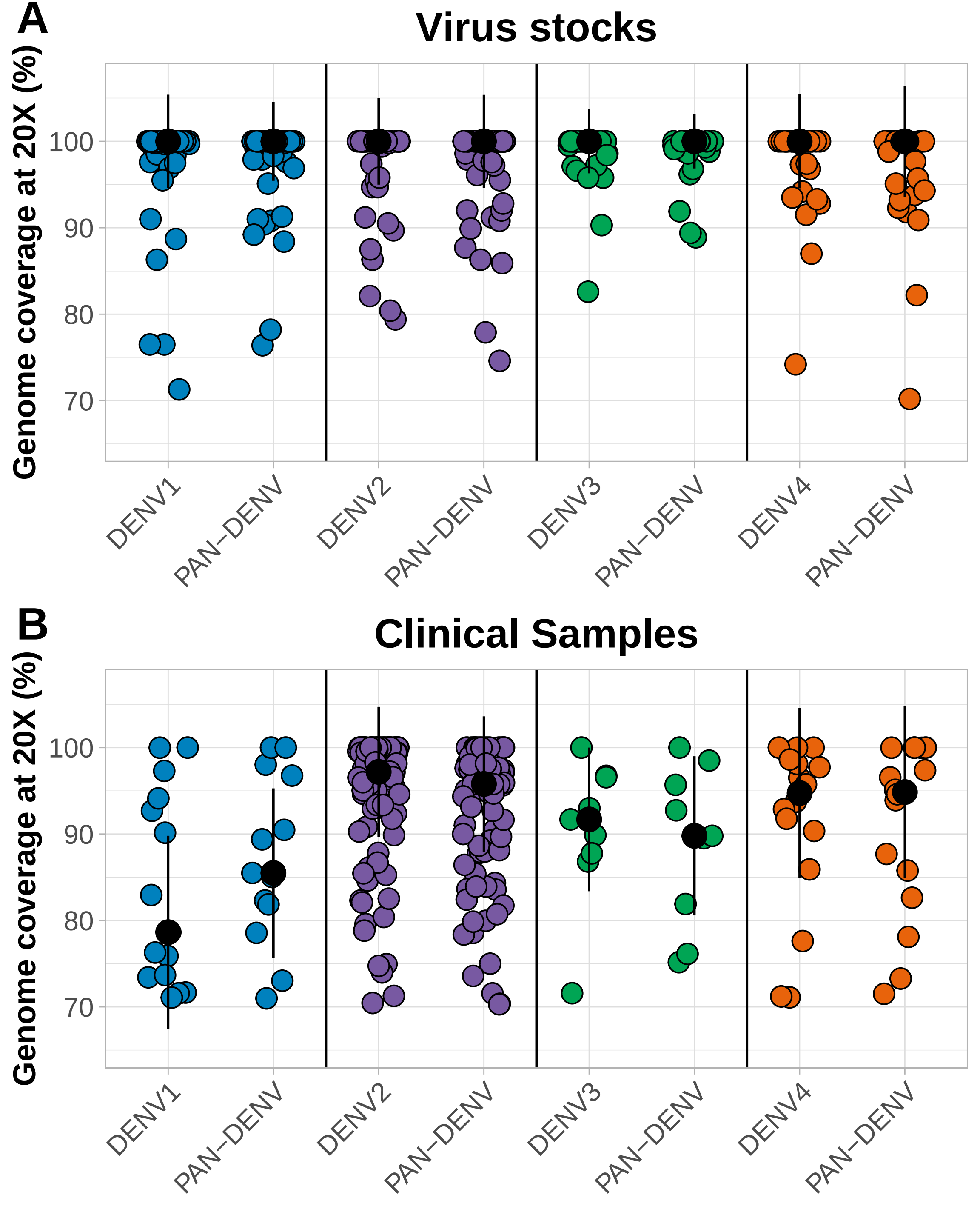
